# Major sex differences in migraine prevalence among occupational categories: a cross-sectional study using UK Biobank

**DOI:** 10.1101/2021.07.27.21261191

**Authors:** Oreste Affatato, Maud Miguet, Helgi B. Schiöth, Jessica Mwinyi

## Abstract

Migraine represents one of the most prevalent neurological conditions worldwide. It is a disabling condition with high impact on the working situation of migraineurs. Interestingly, gender-related differences regarding an association of migraine with important occupational characteristics has been hardly studied. The current study scrutinizes gender-specific differences in the prevalence of migraine across a broad spectrum of occupational categories, shedding also light on associations with important job-related features such as shift work, job satisfaction, and physical activity. The study included data from 415 712 participants from the UK Biobank cohort, using the official ICD10 diagnosis of migraine and other health conditions as selection criteria. Prevalence ratios of migraineurs compared to healthy controls among different occupational categories and job-related variables were estimated using log-binomial regression analyses. Statistical models were adjusted for important sociodemographic features such as age, BMI, ethnicity, education and neuroticism. To better highlight specific differences between men and women we stratified by sex. We detected a differential prevalence pattern in relation to different job categories between men and women. Especially in men, migraine appears to be more prevalent in highly physically demanding occupations (PR 1.38, 95% CI [0.93, 2.04]). Furthermore, migraine is also more prevalent in jobs that frequently involve shift or night shift work compared to healthy controls. Interestingly, this prevalence is especially high in women (shift work PR 1.45, 95% CI [1.14, 1.83], night shift work PR 1.46, 95% CI [0.93, 2.31]). Our results show that higher migraine prevalence is associated with physically demanding jobs and shift working.

## Introduction

Migraine is a highly prevalent neurological disorder that affects predominantly women [1–3]. Due to its highly debilitating symptoms, migraine has an important impact on the daily life of people, interfering negatively with private events, such as time spent with friends and family, as well as with the work productivity. In a study published in 2018 [2], it has been estimated that migraine is the cause of 45.1 million years of healthy life lost due to disability (YLDs) worldwide, where women aged 15 to 49 years are especially affected and account for 20.3 million YLDs. For these reasons, it is of crucial importance to detect the risk factors that can trigger the migraine, in order to develop better preventive strategies. Furthermore, these preliminary epidemiological data show already that it is necessary investigating gender-specific preventive strategies for migraine that take into account the differential prevalence of migraine among men and women.

Many are the risk factors that are associated with migraine. Among the modifiable risk factors, work-related stressors have recently been found to be associated with migraine onset [4]. As a matter of fact, the work may influence the health of a person in many ways, e. g. through working hours, shift or non-shift work, responsibility load, and healthy environment among others. In a prospective cohort study [5] it has been found that a high effort-reward imbalance among women employed in the public sector is associated with an increased risk of a new migraine onset. A longitudinal study of adult health conducted in Brazil [6, 7] showed that low job control, high job demands, low social support, strain-based interference of family with work, and lack of time for personal care and leisure were associated with migraine among general workers. Other risk factors that have been highlighted in recent studies are heavy workloads, emotional stress, shift working and sleep disturbances [8–10]. A cross-sectional study [11] showed also that an ethnic minority with high unemployment rate manifested higher migraine diffusion than a Spanish reference population, especially in women. Accounted risk factors are familiar stress, psychological overload and few hours of sleeping. Another evidence of the relationship between migraine and work-related triggers is a longitudinal study [12], in which the cohort comprised employees in a French gas and electricity company. The aim of this study was to examine the trajectory of migraine in relation with retirement. It has been found that retirement is significantly associated with a decrease in headache prevalence, particularly among subjects with high amount of work stress or with stress-prone personality, such as high hostility or type A personality. A Danish cross-sectional study [13] highlighted more practical issues related to work, like the physical work load and the physical activity. In particular, it has been found that migraine is positively associated with seated/standing and walk jobs. It has been detected also an increased risk to develop migraine in women practicing heavy physical work. From this brief review of the literature it emerges that finding possible correlations between migraine pathogenesis and work-related issues is a complex process, since many interconnected factors may increase the risk for the development of this disorder. Previous studies addressed some of these factors, but a more comprehensive analysis is needed.

A growing body of evidence suggests that work stress may predict a variety of health problems, such as depression, cardiovascular and muscoloskeletal diseases [5, 14–16], but little is known about a possible correlation between migraine and work-related factors [4]. Also, all issues related to work activity are country-specific to a certain extent, and the studies we referred to addressed the association between some work-related issues and migraine, taking into account the cultural and geographical specificity. Therefore, it is important to investigate to what extent these epidemiological findings can be generalized to broader target populations. Also, many studies focused on particular sectors of employment or companies, without addressing the broader issue of the migraine prevalence among the different occupational categories. Another important aspect hardly considered in previous studies are sex-specific differences in relation to job-related features and migraine. This highlights the need for a more precise approach for studying the association between work-related risk factors, sexual gender and migraine, reporting more evidences to clarify some diverging results in the literature [13]. A better understanding of this relationship is crucial to develop more specific and effective preventive strategies, lowering the impact of migraine on society. The aim of this study is therefore to investigate the gender-specific association between occupational categories and work-related features such as physical activity involved, shift work, job satisfaction and migraine, using the large population-based UK Biobank cohort.

## Materials and Methods

### Data source

Phenotype data are provided by the UK Biobank, an ongoing, large population based prospective cohort study in the United Kingdom that began in 2006. The purpose of this biobank is to promote research into the most common and life-threatening diseases. Over half a million volunteers, enrolled at ages from 40 to 75, were invited to visit assessment centers in which they completed questionnaires about their lifestyle and medical history. Basic physical and physiological variables (such as weight, blood pressure) were measured as well. This database includes also in-depth genetic information and neuroimaging measures. The ethical approval for the UK Biobank study has been granted by the North-West Multicenter Research Ethics Committee and the use of these data at our Department was further approved by the Regional Ethics Committee of Uppsala (Sweden).

### Primary outcome variables

In order to investigate the prevalence of migraineurs among the various job categories we used the “Job code at visit” variable, that broadly divides the participants into nine different subgroups, according to the Standard Occupational Classification (SOC2000): managers and senior officials, professional occupations, associate professional and technical occupations, administrative and secretarial occupations, skilled trades occupations, personal service occupations, sales and customer service occupations, process, plant and machine operatives and elementary occupations. To further investigate work-specific characteristics we used variables that describe if the job involves mainly walking or standing, whether it involves heavy manual or physical work, or whether it involves shift or night shift work. To further address the work-related individual situation, we took into account variables that describe work/job satisfaction. For our project we used the values of variables at the baseline, i.e. the first visit of the participants at the centers for the collection of data.

### Covariates

Several basic demographic characteristics were considered as relevant covariates as described in the following. Information about the sex has been acquired from central registry at recruitment, and in some cases it has been updated by the participants. Ethnic background and educational status have been self-reported by the participants in a touch-screen questionnaire. The vast majority of participants was white British, therefore the participants were divided into two categories, i.e. “White” and “Others”. Regarding the educational status, we distinguished just between with or without University/College degree, since in previous studies it has been shown that higher education acts as a protective factor for migraine [17]. Furthermore, we considered the age at recruitment, i.e. the age of the participant when attended an Initial Assessment Centre visit. Since weight can exert an influence on both migraine onset and job-related issues, the BMI was also considered, calculated from height and weight measured during the Initial Assessment Centre visit. Another important variable considered is neuroticism. Work perception may be influenced by personality traits. As highlighted in previous studies [18, 19], negative personality traits have great importance when tackling work-related issues and neurologic disorders, and for this reason it has been taken neuroticism into account.

Covariates such as ethnic background, educational status, age, BMI and neuroticism are associated both with the exposure (job and job-related issues) and the outcome (migraine), therefore they were considered confounders. We adjusted for them in our statistical models, while we stratified for sex. The causal Direct Acyclic Graph in Figure 1 summarizes our choice.

**Figure 1.**
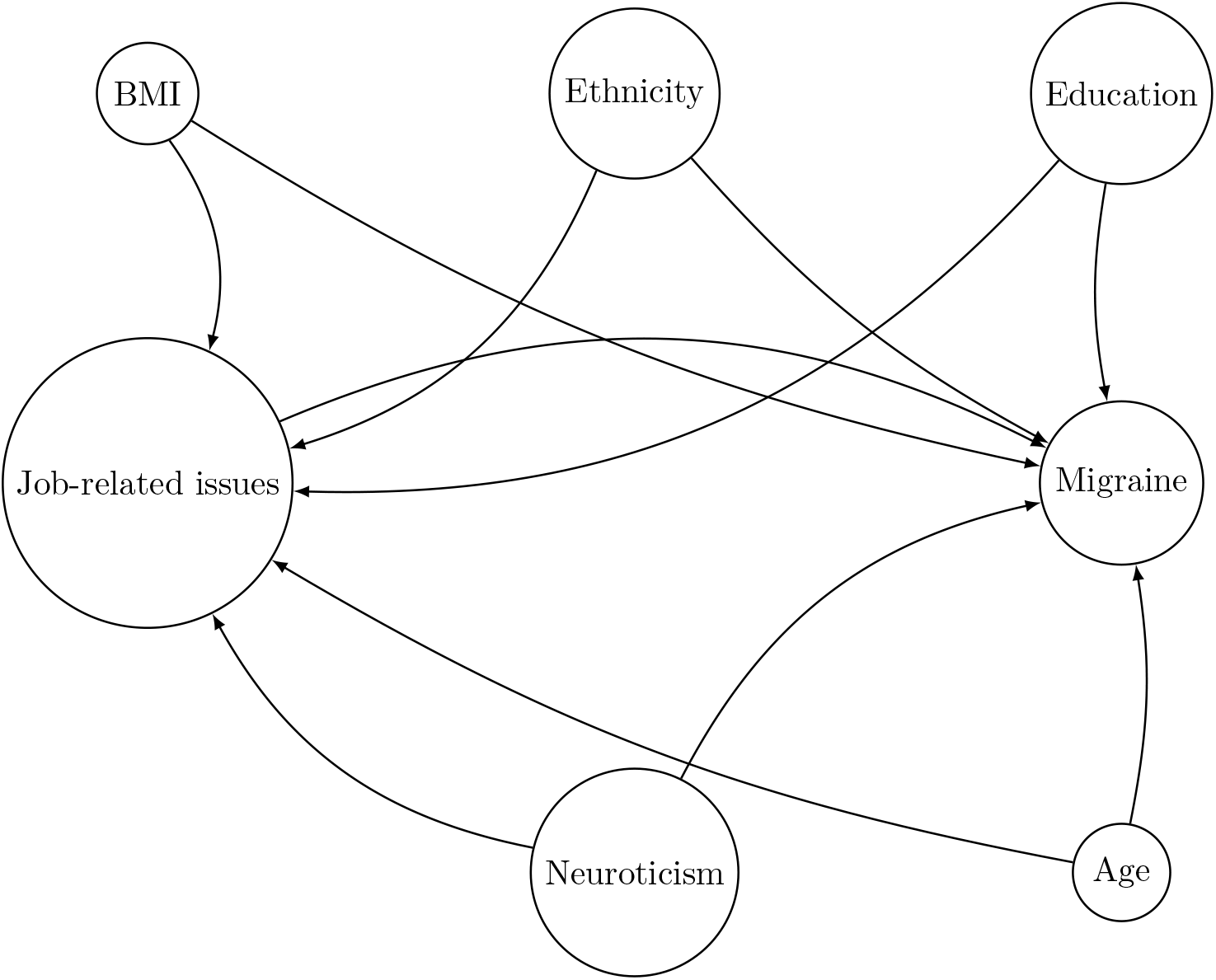
Causal Direct Acyclic Graph illustrating the confounders considered in our model.

### Selection of the participants

The sample initially comprised 502 488 participants. Before proceeding with any analysis, we excluded some participants in virtue of health conditions that may affect our results. The flow chart in Figure 2 summarizes this procedure. Some of the participants decided not to answer to questions about their own health conditions. Those participants were excluded as well, as no sound conclusion could be drawn while including them in the analyses. In any part of the flow chart in which the number of excluded participants was reported, the amount of participants that refused to answer or that were not sure about their own health condition have been included as well.

**Figure 2.**
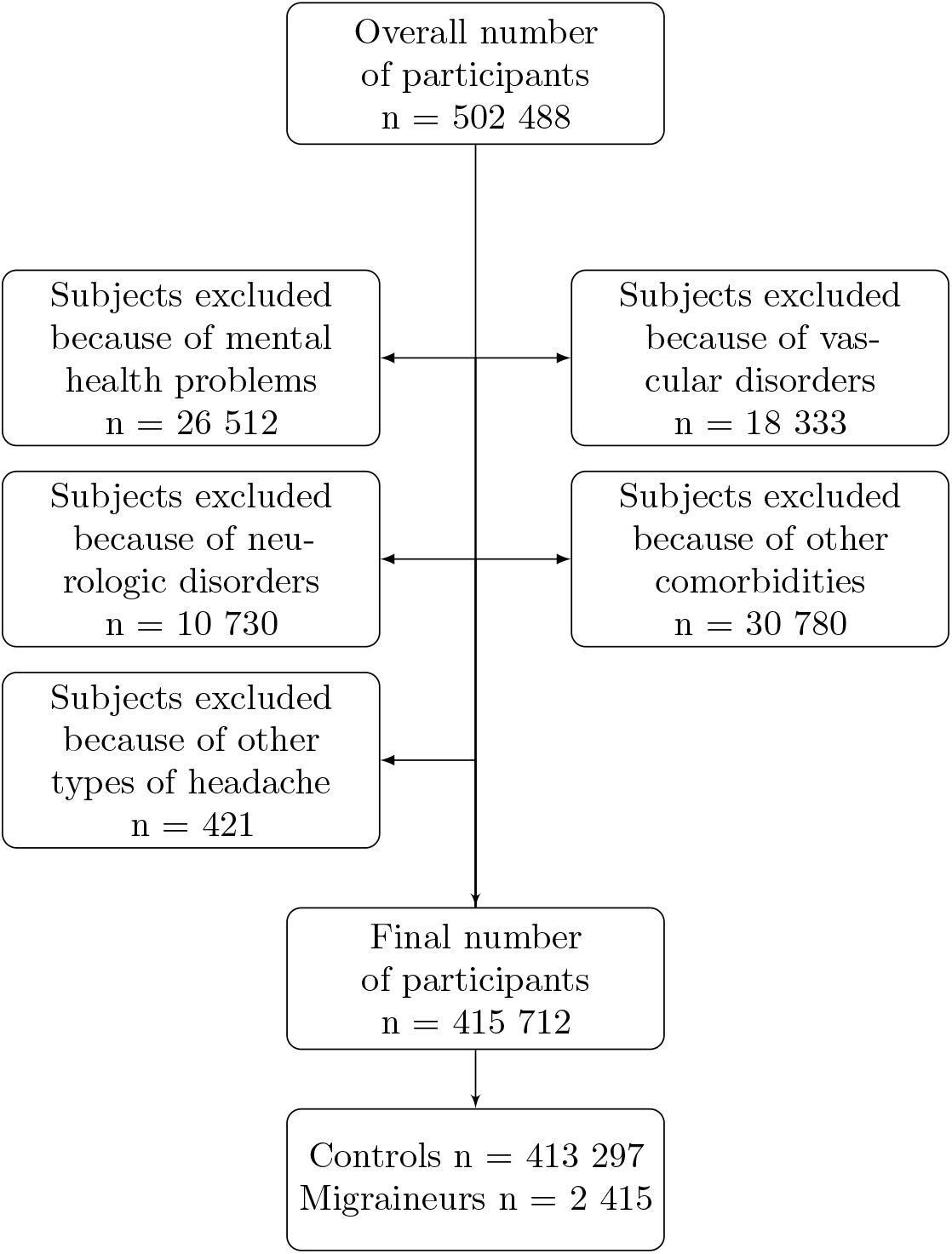
Flow chart of the exclusion process.

The UK Biobank provides information from the hospital registers, ensuring that disorders or illnesses have been at some point diagnosed by a physician. We used the variable “Diagnoses – ICD10” to perform the selection of the participants. Subjects manifesting the most common migraine comorbidities have been excluded, as we wanted to focus our analyses on migraine only, decreasing the interactions with other diseases [20]. We excluded patients with psychiatric comorbidities (bipolar disorder F31, depressive disorder F32-F33, anxiety disorders F41 and post-traumatic stress disorder F431), vascular comorbidities (acute myocardial infarction I21, subsequent myocardial infarction I22, ongoing complications following acute myocardial infarction I23, old myocardial infarction I252, stroke I64, sequelae of stroke I694, brain stem stroke syndrome G463, cerebellar stroke syndrome G464 and Raynaud’s syndrome I730), neurologic comorbidities (epilepsy G40, status epilepticus G41, multiple sclerosis G35, nonorganic sleep disorders F51 and sleep disorders G47), fibromyalgia M797, temporomandibular joint disorder K076, vasomotor and allergic rhinitis J30, asthma J45, status asthmaticus J46, systemic lupus erythematosus M32. In order to achieve a more precise analysis on migraine, we excluded participants with general headaches (other headache syndromes G44) as well.

### Statistical analysis

In order to investigate the prevalence of migraineurs in the various job categories as compared to healthy controls, we calculated the prevalence ratio (PR) with 95% confidence interval (CI). We performed the calculations for PR and 95% CI using two models: (i) adjusting for sociodemographic confounders such as age, BMI, ethnic background (ii) as model (i) including education level and neuroticism as well [21]. The analysis has been stratified by sex, to further investigate the differences between men and women. We performed log-binomial regression analysis for the two models [22, 23]. In three cases it was not possible to perform the analysis for the first model: occupational category and heavy manual or physical work for women and shift work for men. Convergence criteria were not met as the Hessian matrix was singular. In these three cases we added the variable neuroticism as predictor in the model. Only in these three cases the first model had this change, while the fully adjusted model is equal to the others.

As recommended by leading epidemiologists and statisticians, a thorough discussion of effect sizes and confidence intervals has been provided, as the most solid way to draw sensible conclusion from statistical analyses combined with biological scientific background. For these reasons, no statistical test has been performed nor any threshold of statistical significance has been established. P-values have not been provided as possibly misleading [24–32]. Accordingly, any finding deemed as significant is intended to be clinically or epidemiologically (and not statistically) significant.

The statistical analysis has been conducted via SPSS (IBM SPSS Statistics version 26). Plots and diagrams have been created using GraphPad Prism 9.12 and LATEX.

## Results

The flow chart in Figure 2 summarizes the selection process of individuals finally included in the analysis. From the initial sample population of 502 488 participants, 86 776 subjects were excluded (17.3% of the initial sample) as they have been diagnosed by a disease or disorder as listed in the exclusion criteria or because they refused to report their health condition. The final sample was divided into two main categories, i.e. migraineurs and controls. Table 1 shows the main demographic and socioeconomic characteristics of the final sample population. Analysis of data reported in Table 1 allows detecting interesting features. Migraine is more prevalent in women rather than in men, with a 3:1 ratio, in line with what is already known from an epidemiological perspective [1]. The neuroticism score on average is higher in migraineurs as compared with controls. Subjects with high neuroticism score tend to respond worse to stressors and this feature may play an important role in migraine onset, as stress and poor stress response are common risk factors for this neurologic disorder [17]. In general, we can also observe that this sample population is constituted mostly by middle aged, slightly overweight, white British people, whereby variables such age, BMI and education level are well balanced in the study arms.

**Table 1.** Socioeconomic and demographic characteristics of the population.

|  | **Migraineurs**<br>n = 2 415 | **Controls**<br>n = 413 297 |
| --- | --- | --- |
| **Sex** |  |  |
| Women | 1 821 (75.4%) | 225 886 (54.7%) |
| Men | 594 (24.6%) | 187 411 (45.3%) |
| **Age** (mean ± SD) | 55 ± 8 | 56 ± 8 |
| **Ethnic background** |  |  |
| White | 2 276 (94.2%) | 389 051 (94.1%) |
| Others | 139 (5.8%) | 24 246 (5.9%) |
| **Education** |  |  |
| College or University degree | 726 (30.1%) | 138 477 (33.5%) |
| Others | 1 689 (69.9%) | 274 820 (66.5%) |
| **BMI** (mean ± SD) | 27.2 ± 4.8 | 27.2 ± 4.6 |
| **Neuroticism score** (mean ± SD) | 4.76 ± 3.26 | 3.93 ± 3.18 |

Table 2 reports the descriptive statistics of job-related outcome variables. Since not all the selected participants underwent all the various questionnaires, at the beginning of each main category it has been reported the overall number of migraineurs and controls who actually answered that particular job-related questionnaires. Data on occupational categories are available from 1 569 migraineurs and 277 241 controls. Female migraineurs appear to be more prevalent in associate professional and technical occupations, personal services occupations and sales and customer service occupations. In contrast to this, male migraineurs appear to be less prevalent in categories such as managers and senior officials and professional occupations, while they appear to be more prevalent in physically demanding jobs such as the ones included in elementary occupations or in skilled trades occupations.

**Table 2.** Descriptive statistics of job-related variables.

|  | **Migraineurs** |  | **Controls** |  |
| --- | --- | --- | --- | --- |
|  | Women | Men | Women | Men |
| **Job** | **1 157** | **412** | **145 331** | **131 910** |
| Managers and Senior Officials | 147 (12.7%) | 75 (18.2%) | 18 482 (12.7%) | 29 954 (22.7%) |
| Professional Occupations | 234 (20.2%) | 95 (23.1%) | 32 052 (22.1%) | 33 462 (25.4%) |
| Associate Professional,<br>Technical Occupations | 241 (20.8%) | 69 (16.7%) | 28 529 (19.6%) | 19 592 (14.9%) |
| Administrative and<br>Secretarial Occupations | 252 (21.8%) | 31 (7.5%) | 35 122 (24.2%) | 8 197 (6.2%) |
| Skilled Trades Occupations | 11 (0.6%) | 70 (17.0%) | 2 451 (1.7%) | 17 908 (13.6%) |
| Personal Service Occupations | 143 (12.4%) | 10 (2.4%) | 13 675 (9.4%) | 2 726 (2.1%) |
| Sales and Customer Service Occupations | 66 (5.7%) | 4 (1.0%) | 7 098 (4.9%) | 2 249 (1.7%) |
| Process, Plant and Machine Operatives | 13 (0.7%) | 27 (6.6%) | 1 558 (0.7%) | 10 702 (8.1%) |
| Elementary Occupations | 50 (4.3%) | 31 (7.5%) | 6 364 (4.4%) | 71 20 (5.4%) |
| **Heavy manual or physical work** | **1 048** | **362** | **127 964** | **118 577** |
| Never/rarely | 719 (68.6%) | 207 (57.2%) | 90 197 (70.5%) | 71 449 (60.3%) |
| Sometimes | 225 (21.5%) | 75 (20.7%) | 25 591 (20.0%) | 26 867 (22.7%) |
| Usually | 55 (5.2%) | 39 (10.8%) | 6 226 (4.9%) | 10 258 (8.7%) |
| Always | 49 (4.7%) | 41 (11.3%) | 5 950 (4.6%) | 10 003 (8.4%) |
| **Work involves mainly**<br>**walking/standing** | **1 048** | **362** | **127 982** | **118 510** |
| Never/rarely | 362 (34.5%) | 115 (31.8%) | 48 128 (37.6%) | 39 967 (33.7%) |
| Sometimes | 294 (28.1%) | 103 (28.5%) | 37 579 (29.4%) | 37 906 (32.0%) |
| Usually | 173 (16.5%) | 56 (15.5%) | 18 235 (14.2%) | 17 965 (15.2%) |
| Always | 219 (20.9%) | 88 (24.3%) | 24 040 (18.8%) | 22 672 (19.1%) |
| **Job involves shift work** | **1 045** | **360** | **127 815** | **118 403** |
| Never/rarely | 865 (82.8%) | 288 (80.0%) | 109 261 (85.5%) | 95 321 (80.5%) |
| Sometimes | 70 (6.7%) | 31 (8.6%) | 7 940 (6.2%) | 10 031 (8.5%) |
| Usually | 20 (1.9%) | 7 (1.9%) | 2 459 (1.9%) | 2 697 (2.3%) |
| Always | 90 (8.6%) | 34 (9.4%) | 8 155 (6.4%) | 10 354 (8.7%) |
| **Job involves night shift work** | **182** | **73** | **18 737** | **23 212** |
| Never/rarely | 101 (55.5%) | 30 (41.1%) | 10 711 (57.2%) | 9 805 (42.2%) |
| Sometimes | 41 (22.5%) | 25 (34.2%) | 4 569 (24.4%) | 7 497 (32.3%) |
| Usually | 12 (6.6%) | 7 (9.6%) | 1 204 (6.4%) | 2 188 (9.4%) |
| Always | 28 (15.4%) | 11 (15.1%) | 2 253 (12.0%) | 3 722 (16.0%) |
| **Job satisfaction** | **425** | **142** | **54 613** | **47 353** |
| Extremely happy | 37 (8.7%) | 13 (9.2%) | 4 305 (7.9%) | 4 151 (8.8%) |
| Very happy | 155 (36.5%) | 51 (35.9%) | 20 053 (36.7%) | 16 993 (35.9%) |
| Moderately happy | 200 (47.1%) | 57 (40.1%) | 25 785 (47.2%) | 20 968 (44.3%) |
| Moderately unhappy | 24 (5.6%) | 13 (9.2%) | 3 354 (6.1%) | 3 818 (8.1%) |
| Very unhappy | 6 (1.4%) | 4 (2.8%) | 757 (1.4%) | 1 017 (2.1%) |
| Extremely unhappy | 3 (0.2%) | 4 (2.8%) | 359 (0.7%) | 406 (0.9%) |

To assess the influence of job-related physical strain on migraine, we considered the variables in our analyses that described the frequency of heavy manual or physical work as well as the intensity of walking or standing during work. In the first case, 1 410 migraineurs and 246 541 controls completed the questionnaire while in the second case it was completed by 1 410 migraineurs and 246 492 controls. In general, migraine is more prevalent in jobs requiring constant demanding physical activity.

Another feature of the job categories that we addressed in relation to migraine is shift work. The questionnaire about shift work was completed by 1 405 migraineurs and 246 218 controls, while the one about night shift work was completed by 255 migraineurs and 41 949 controls. From the data we can see that migraineurs are slightly more prevalent in jobs that involve constant shift or night shift work, and this holds true especially for women.

### Statistical analysis

Table 3 and Figure 3 display prevalence ratios (PR) and corresponding 95% confidence intervals (95% CI) for the different occupational categories, as classified according to SOC2000, and stratified according to the sex of the participants. Although point estimates of the PRs are limited in the precision because of wide 95% CI (see Figure 3), the following observations can be made. Male migraineurs are less prevalent in managerial (fully adjusted PR 0.50, 95% CI [0.31, 0.80]), professional (fully adjusted PR 0.60, 95% CI [0.38, 0.95]) and associate professional occupations (fully adjusted PR 0.70, 95% CI [0.44, 1.13]). Female migraineurs tend to be less prevalent in skilled trades (fully adjusted PR 0.58, 95% CI [0.28, 1.20]) and administrative and secretarial occupations (fully adjusted PR 0.86, 95% CI [0.61, 1.21]).

**Table 3.**
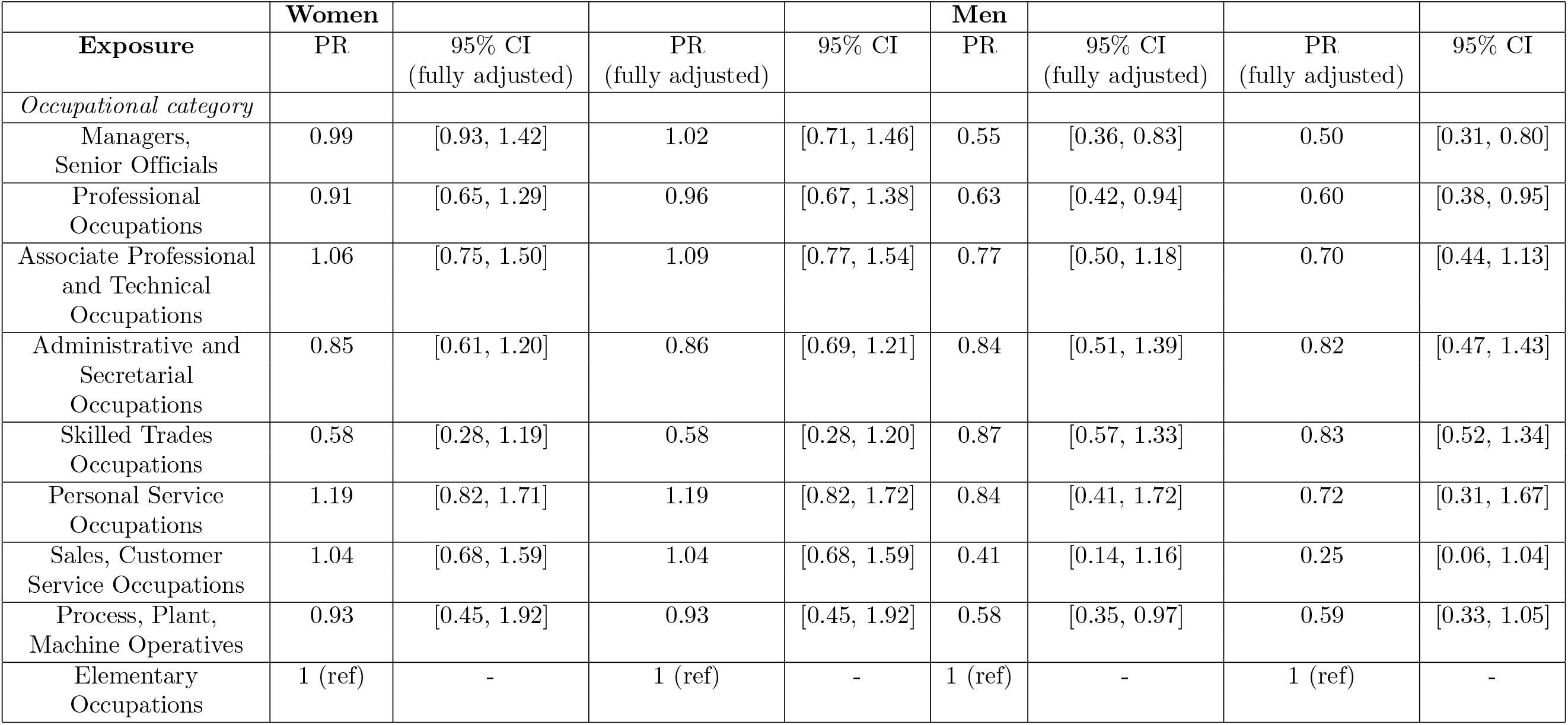
Prevalence of migraineurs among different occupational categories.

**Figure 3.**
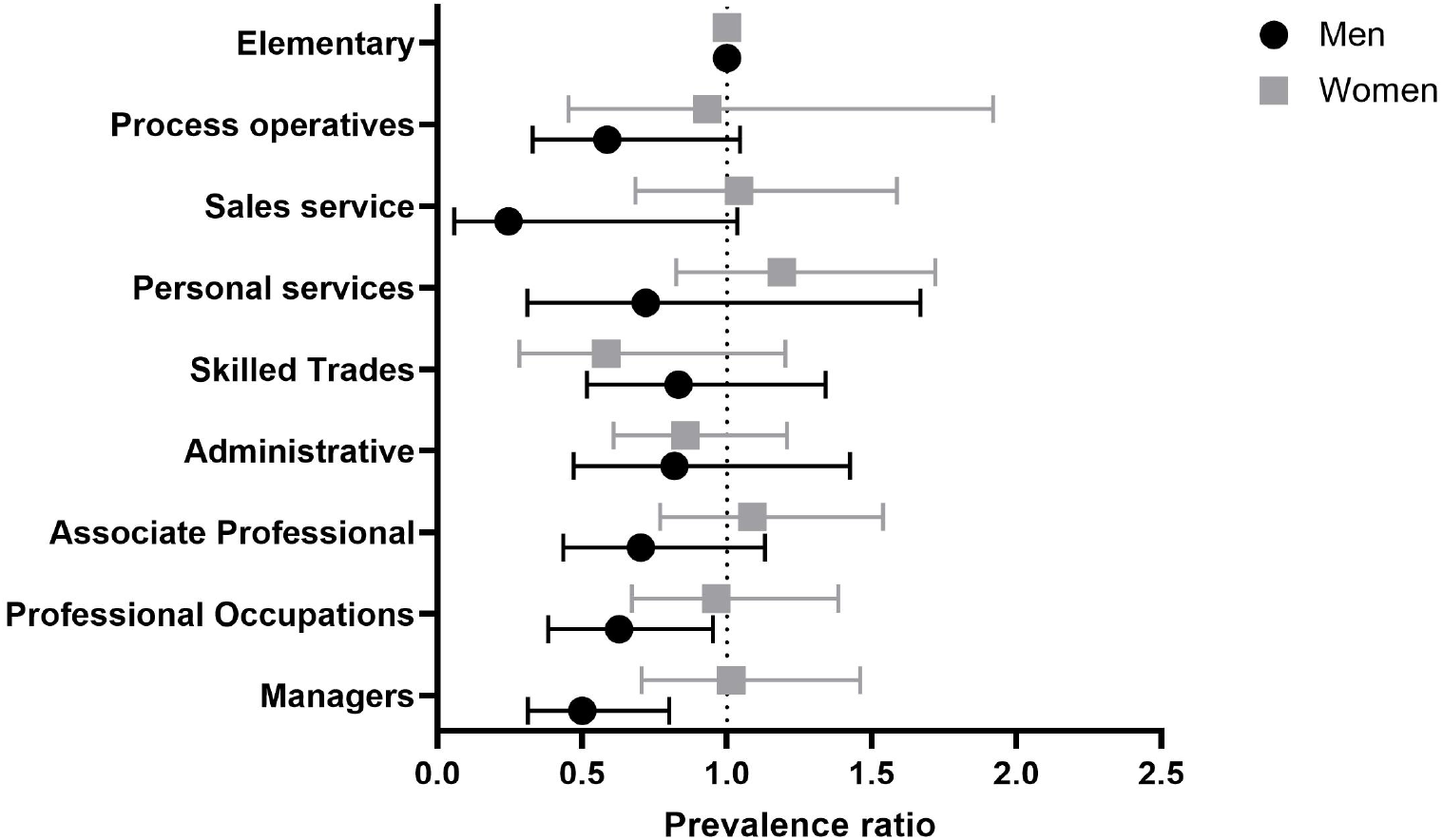
Forest plot illustrating the prevalence of migraine among occupational categories. Means with error bars plot of the data from Table 3. The point estimate is the prevalence ratio (fully adjusted model) of migraineurs compared with controls in the different occupational sectors as defined by SOC2000.

**Figure 4.**
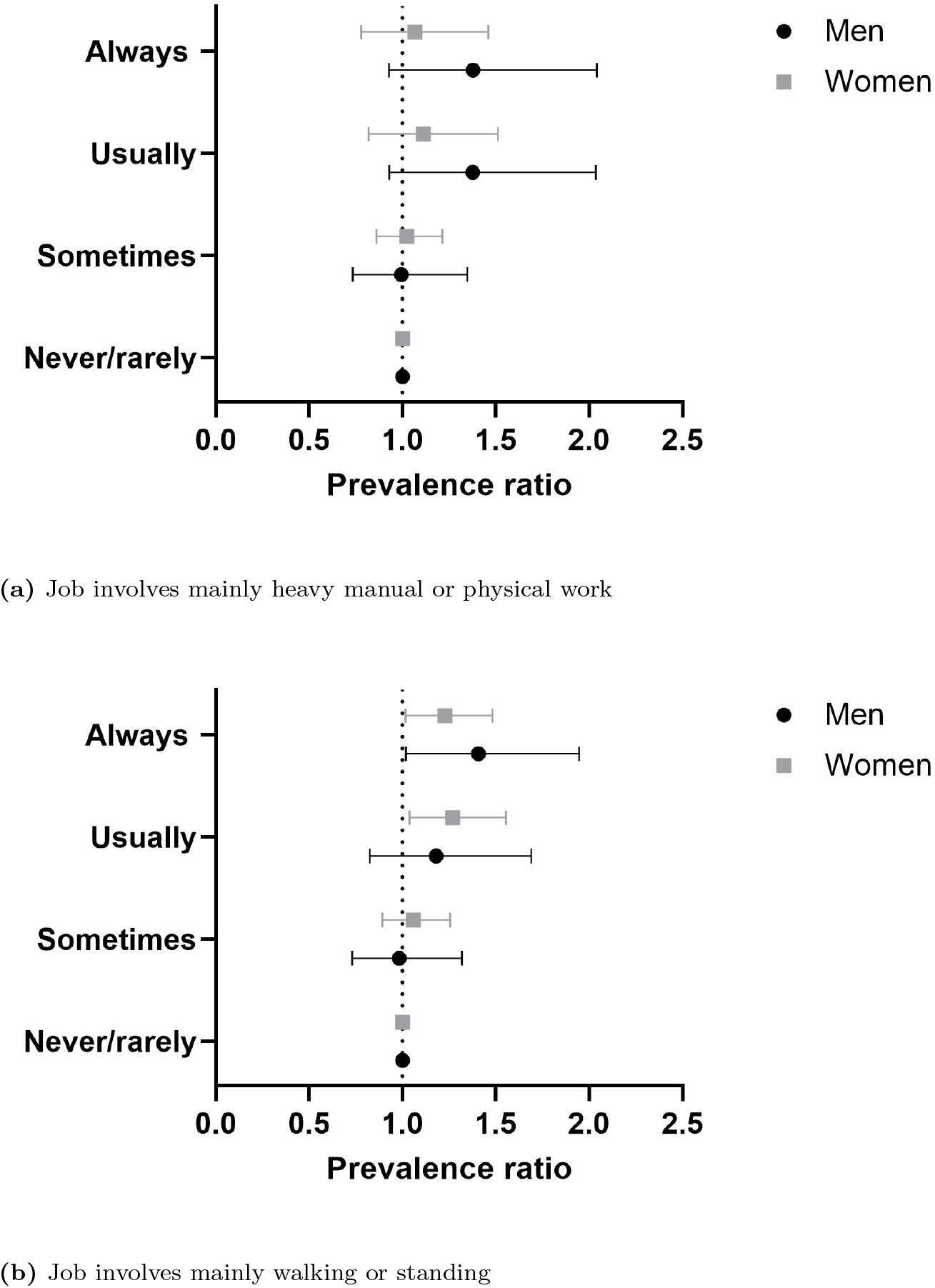
Forest plot illustrating the prevalence of migraine among different occupational categories, divided by work-related physical activity. Mean with error bars plots of the data from Table 4. The point estimate is the prevalence ratio (fully adjusted model) of migraineurs compared with controls among job categories, differentiated by the relevance and frequency of (a) heavy manual or physical work and (b) walking or standing.

**Figure 5.**
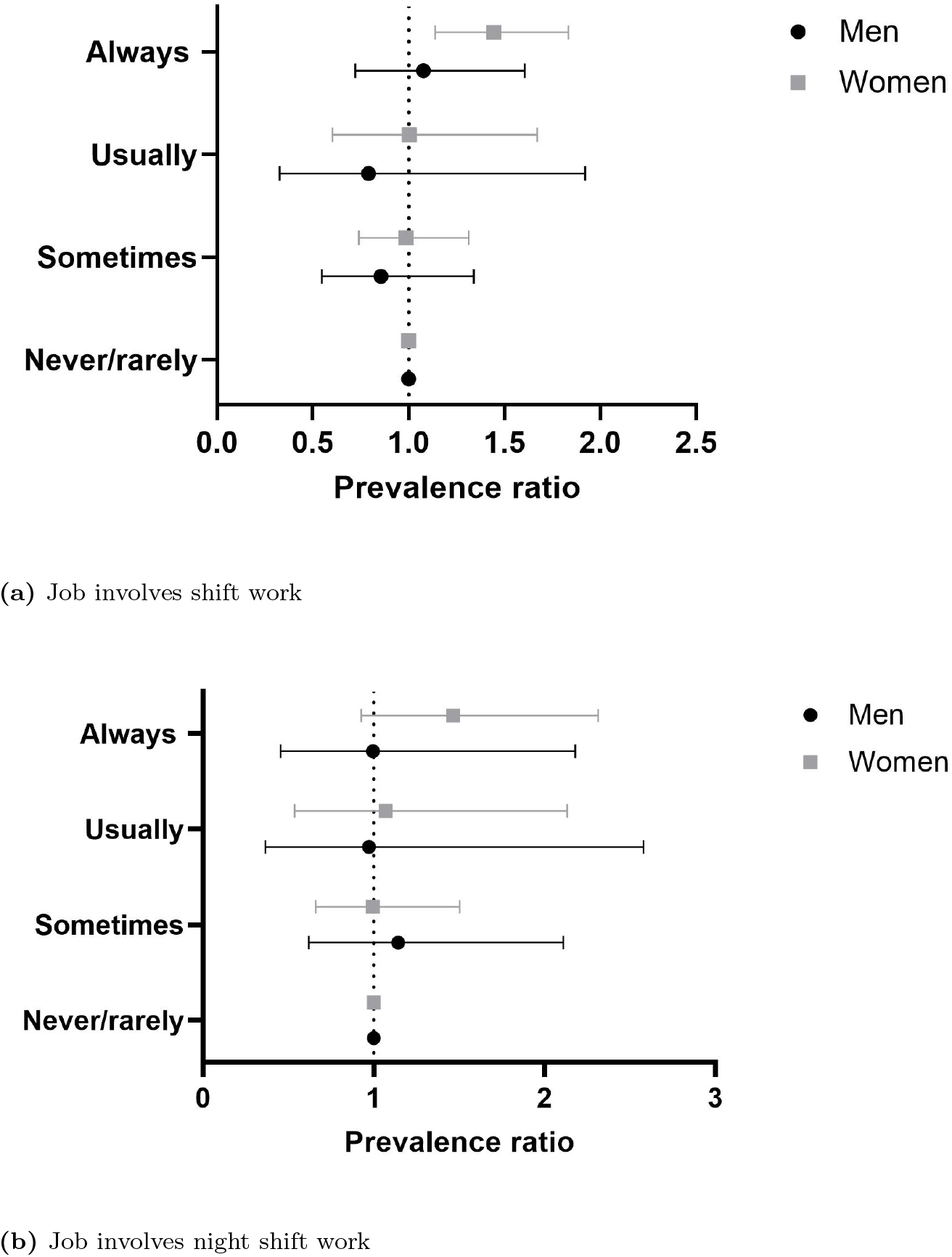
Forest plot illustrating the prevalence of migraine among different occupational categories, divided by the type of shift work. Mean with error bars plots of the data from table 5. The point estimate is the prevalence ratio (fully adjusted model) of migraineurs compared with controls among job categories, differentiated by frequency of (a) shift work and (b) night shift work.

Table 4 shows prevalence ratios and 95% confidence intervals for different frequency or intensity levels of exposure to heavy physical work and walking/standing at work. Migraine results to be more prevalent in men that are more often involved in heavy manual or physical work (fully adjusted PR 1.38, 95% CI [0.93, 2.04]) and in walking or standing (fully adjusted PR 1.41, 95% CI [1.02, 1.95]). The same feature is shared by women, with higher prevalence of migraine among participants with jobs characterized by frequent walking/standing (fully adjusted PR 1.23, 95% CI [1.02, 1.48]).

**Table 4.**
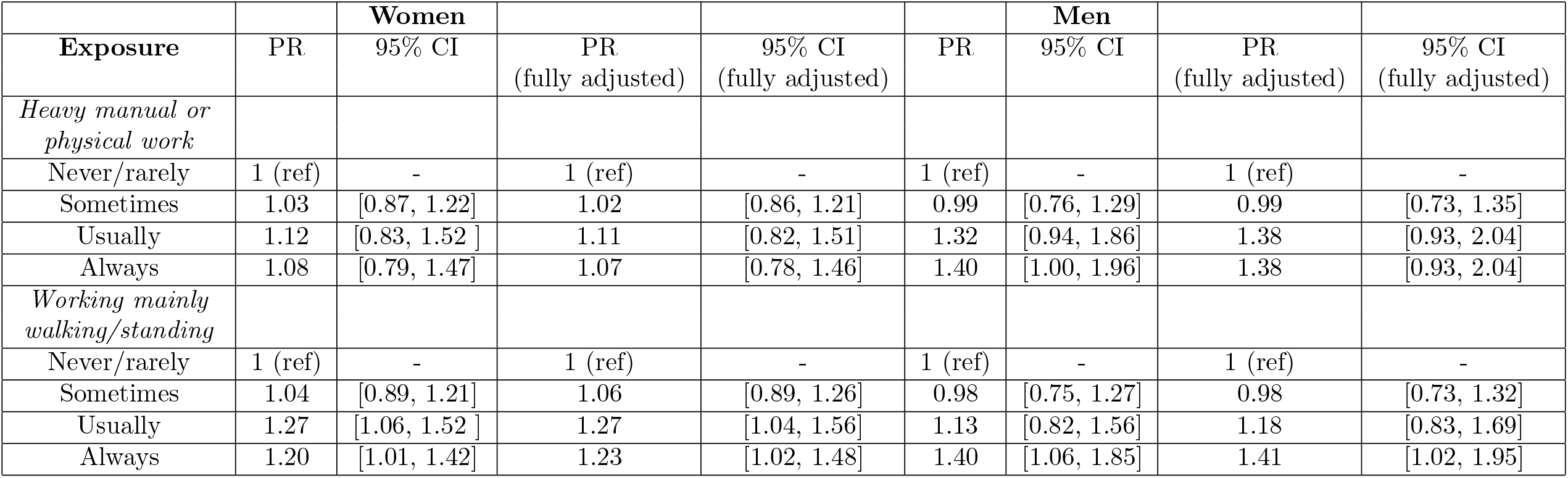
Prevalence of migraineurs among different job-related categories (physical activity).

Table 5 shows the results of our analyses investigating the prevalence of migraine among different kinds of shift works. Female migraineurs are more prevalent among jobs that involve shift work (fully adjusted PR 1.45, 95% CI [1.14, 1.83]). A similar trend is observed for night shift work (fully adjusted PR 1.46, 95% CI [0.93, 2.31]), although 95% CI are in general wide due to low number of participants, thus limiting the possibility to draw robust conclusions.

**Table 5.** Prevalence of migraineurs among different job-related categories (shift work).

| **Exposure** | **PR** | **Women**<br>95% CI | **PR**<br>(fully adjusted) | **95% CI**<br>(fully adjusted) | **PR1** | **Men**<br>95% CI | **PR**<br>(fully adjusted) | **95% CI**<br>(fully adjusted) |
| --- | --- | --- | --- | --- | --- | --- | --- | --- |
| *Job involves shift work* |  |  |  |  |  |  |  |  |
| Never/rarely | 1 (ref) | - | 1 (ref) | - | 1 (ref) | - | 1 (ref) | - |
| Sometimes | 1.09 | [0.85, 1.39] | 0.99 | [0.74, 1.31] | 0.88 | [0.57, 1.38] | 0.86 | [0.55, 1.34] |
| Usually | 1.02 | [0.66, 1.59] | 1.00 | [0.60, 1.67] | 0.83 | [0.34, 2.01] | 0.79 | [0.33, 1.92] |
| Always | 1.35 | [1.08, 1.68] | 1.45 | [1.14, 1.83] | 1.142 | [0.77, 1.69] | 1.08 | [0.72, 1.61] |
| *Work involves night shift work* |  |  |  |  |  |  |  |  |
| Never/rarely | 1 (ref) | - | 1 (ref) | - | 1 (ref) | - | 1 (ref) | - |
| Sometimes | 0.94 | [0.65, 1.35] | 0.99 | [0.66, 1.50] | 1.02 | [0.60, 1.73] | 1.14 | [0.62, 2.11] |
| Usually | 1.07 | [0.59, 1.95] | 1.07 | [0.54, 2.13] | 0.96 | [0.42, 2.19] | 0.97 | [0.37, 2.58] |
| Always | 1.33 | [0.88, 2.03] | 1.46 | [0.93, 2.31] | 0.90 | [0.45, 1.80] | 1.00 | [0.45, 2.18] |

Table 6 and Figure 6 display the outcomes of our analyses on the relationship between prevalence of migraine and job satisfaction. Participants were asked to rate how satisfied they were with the work they were doing. The general trend for both man and women is that migraine is less prevalent among participants happier with their job. In particular, migraine appears to be less prevalent among women moderately happy with their job (fully adjusted PR 0.76, 95% CI [0.52, 1.12]). Men belonging to the “extremely unhappy” group show higher prevalence of migraine, but being the 95% CI very wide it is difficult to draw a solid conclusion on the actual size of this effect.

**Table 6.**
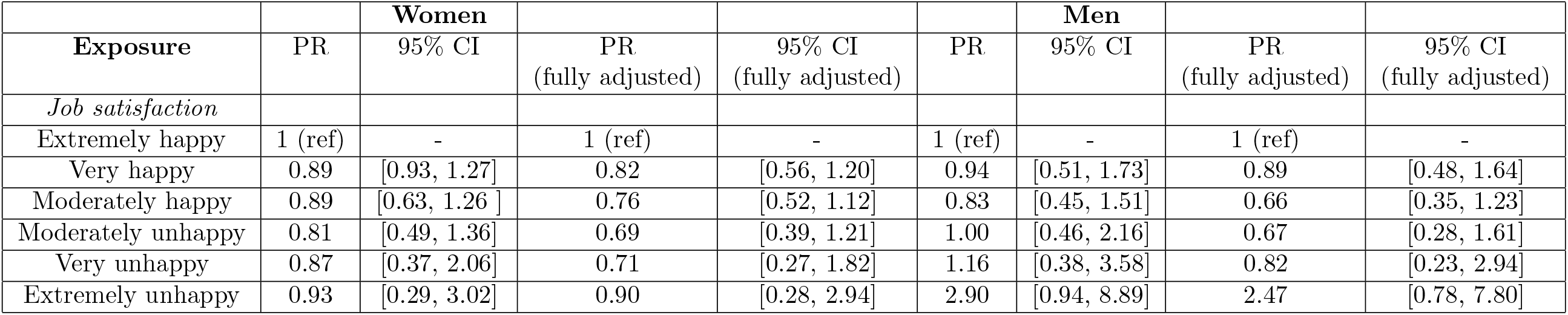
Prevalence of migraineurs among different job-related categories (work satisfaction).

**Figure 6.**
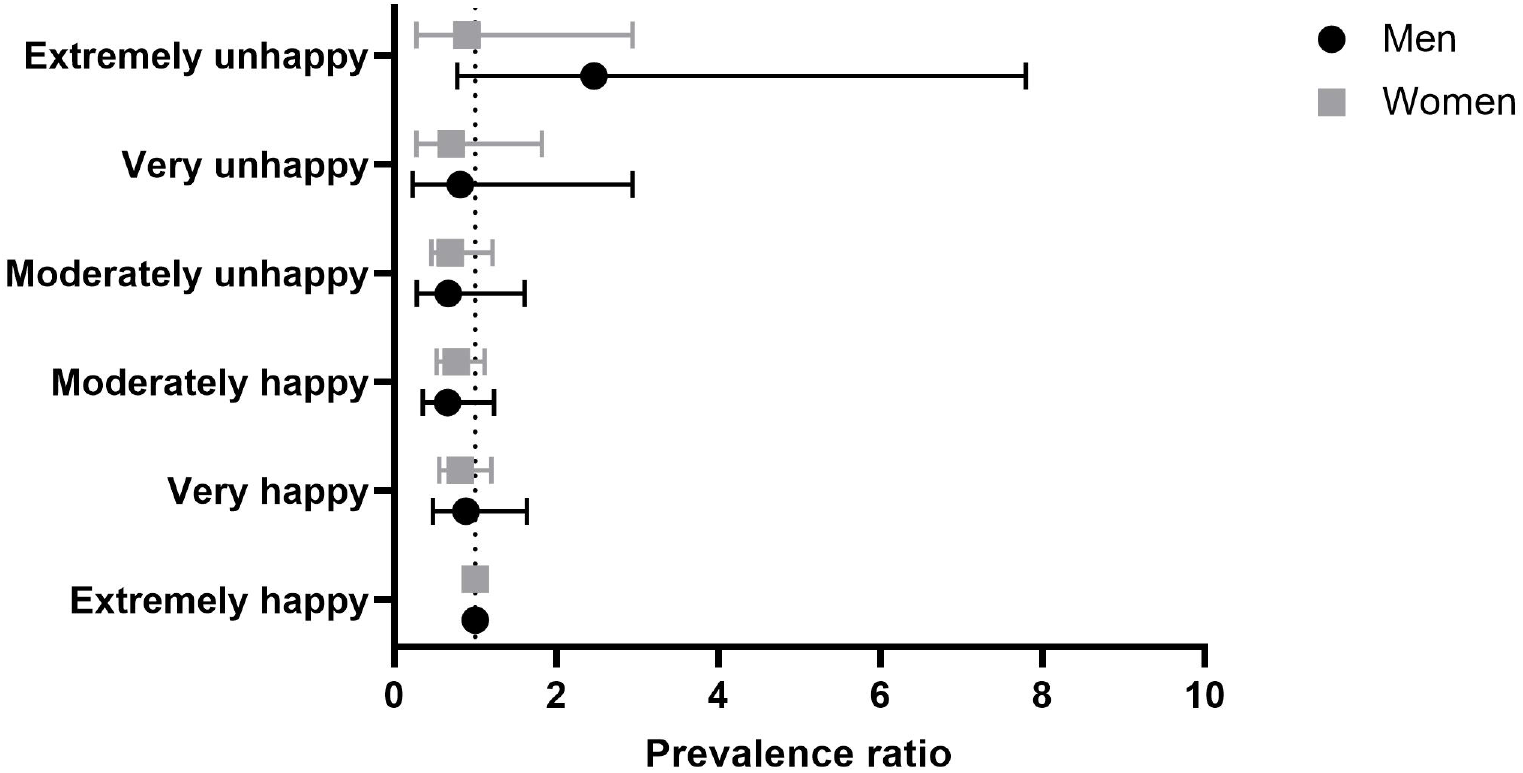
Forest plot illustrating the prevalence of migraine among participants divided by different degrees of job satisfaction. Mean with error bars plots of the data from table 6. The point estimate is the prevalence ratio (fully adjusted model) of migraineurs compared with controls among different levels of job satisfaction.

## Discussion

To our knowledge, this is the first study that thoroughly addresses the prevalence of migraine in a broader spectrum of occupational categories. While other studies focused only on a very particular work sector (e.g. health professionals) [5–12], we considered nine different job categories (according to SOC2000) as well as important job related features such as work-related physical activity, shift work and job satisfaction. Furthermore, stratifying by sex we were able to perform more precise analyses, highlighting sex-specific differences on the prevalence pattern of migraine among the various job-related features. The fact that we considered official diagnosis from hospital registry is another important feature of this study, as it is a more solid and trustworthy way to divide patients and controls as compared with analyses based on self-reported diagnosis.

Our results show striking differences between men and women in the prevalence of migraine dependent on occupational categories. Overall, in women the prevalence of migraine is not significantly different from healthy controls, across the different jobs. Of note, in two categories, namely administrative and secretarial occupations and skilled trade occupations, female migraineurs are less prevalent than female controls. The first category comprises people working as administrative officers, assistants and clerks in government and related organizations, finance, records and communications as well as secretarial occupations in various sectors (e.g. medical, legal and education areas). The second one comprises people working in agricultural, metal, electrical, construction, textiles and printing trades. Administrative and related occupations may be mentally and psychologically demanding, while the trade-related occupations may be rather physically demanding. Thus, both may be a significant source of stress, which is a risk factor for migraine [3, 4, 20]. The fact the migraine is not highly prevalent in these groups is probably due to the fact that these types of job are also characterized by a high level of control and regulation. This may act as a protective factor against the onset of migraine, as it has been shown in a previous study that low control in the work environment is a risk factor [6]. This argument is supported by our analyses on work satisfaction, showing that a good work environment is inversely associated with migraine.

For men the situation appears to be the opposite. Physically demanding occupations are characterized by a higher prevalence of migraineurs, while the others, more psychologically demanding, have a lower prevalence. This is also confirmed by the analyses on work-related physical activity, as discussed below. In particular, managers and senior, professional, associate professional and technical occupations are characterized by very low prevalence of migraine. These occupational categories comprise many different types of job, such as managers in all sectors, science and technology, health, teaching and research, business and public service professionals as well as assistants and technical occupations related to these areas. These jobs require highly educated and skilled workers. Thus, it can be hypothesized that they may have a better access to healthier environments and therapeutic interventions that can improve the health in general and reduce the risk of migraine. However, this is also true for women, so this cannot explain the asymmetry between the prevalence of migraine for men and women in this various job categories. Additional sex-specific mechanisms may act to account for this asymmetry. With the data available for this research we were not able to further address this issue.

The analyses on the work-related physical activity show that frequent heavy manual physical work is related to higher prevalence of migraine among men, while for women there is no significant difference compared with the healthy controls. This interaction between migraine and physical activity is also supported by the analyses on jobs that involve mainly walking or standing. Higher prevalence of migraine has been reported among men and women for whom it’s required to walk or stand for many hours and often at their work place. There is no conclusive evidence in the scientific literature about the impact of physical exercise on migraine. Is it a risk or a protective factor? A recent review [31] addressed the issue of the relation between migraine and physical exercise. There is a limited amount of evidence that proves that intensive physical exercise may trigger migraine attacks, however there are extensive studies that prove that regular aerobic exercise may be an effective preventive strategy against migraine. In general, the review reported studies on regular and intensive physical exercise, not related to the work place, where the intensity of the physical activity and the duration may vary consistently. It could be possible that these irregular working rhythms are the source of a kind of stress that may be at the origin on migraine onset for men. Another feature related to these elementary occupations that should be taken into account is the general lifestyle of these workers, as many habits and sociodemographic factors can increase the migraine prevalence (e.g. obesity, caffeine consumption among others) [3].

The level of the impact of shift work on migraine has not been fully established, as the available data are few and conflicting [9, 32]. One key argument in favor of the possible migraine risk due to shift work is the variation of the circadian rhythm. Shift and night shift work are characterized by irregular schedule and this affects negatively the sleep-wake balance and thus making irregular the circadian rhythm. This may be at the origin of migraine pathogenesis. Our analyses show that migraine is more prevalent in jobs characterized by frequent shift and night shift work. In particular, the prevalence is higher for women and for night shift work, supporting the sleep-wake imbalance theory as the possible explanation of the causing factor of migraine. Not many participants answered the shift work questionnaires, thus our analyses are characterized by lower precision, due to the smaller sample sizes, as compared with previous analyses.

Some limitations of this study derives from some features of the UK Biobank itself. The sample population is characterized by older people (the mean is around 56 with a standard deviation of 8 years), while migraine is mostly prevalent in people between 35 and 39 [1]. Also, not all the participants answered all the work-related questionnaires, and this forced us to perform analyses with different degrees of precision, due to the variation in sample sizes. Another limitation is due to the fact that there is no data available on when the participants were actually diagnosed with migraine (or any other condition considered for the selection of the participants). In particular, some participants may have been not affected by migraine when their data have been collected. One last limitation of this project is its cross-sectional nature. This allowed us to give an estimation of the prevalence of migraine among different work-related groups, but not to assess risk or incidence. Any kind of hypothesis about the possible underlying causative path between the examined exposures and the onset of migraine in this study is purely speculative, even though supported by parallel and independent evidence from other studies. Nonetheless, the study of the prevalence of migraine among the different groups here considered is a powerful tool that can be used by policy makers and health institutions to improve the work environment and therefore the health of the workers.

## Conclusion

Previous studies showed how particular job-related features exert a differential impact on women and men in terms of migraine pathogenesis. This research shows that migraine is more prevalent in physically demanding occupations, especially in men. Migraine is also more prevalent in jobs that involve frequently shift or night shift work, and the prevalence is even higher in women. Study results support the assumption of gender-specific differences in work-related stressors associated to migraine.

## Data Availability

The data that supports the findings of this project are available from UK Biobank. All data generated during this study are included in this paper.

## Declarations

### Authors’ contributions

JM and OA conceived and designed the study. JM, OA and MM developed the methodology hereby adopted. OA performed the formal analysis, took care of the data curation and wrote the original draft. JM, MM, and HBS supervised the project. All the authors reviewed and edited the manuscript.

### Authors’ approval

All authors have seen and approved the manuscript.

### Ethics approval

We declare to have ethical permission to use UK Biobank data (UKB permission UKB 57519).

## Acknowledgments

We thank all the participants for their support in this research.

